# Sex-stratified analysis of the potential association between *PGLYRP2* rs892145 variant and Parkinson’s disease across diverse ancestral populations

**DOI:** 10.1101/2025.07.23.25331993

**Authors:** Cesar Luis Avila, Henry Mauricio Chaparro-Solano, Valentina Quintana-Peña, Kajsalisa Åberg, Kajsa Atterling Brolin, Global Parkinson’s Genetics Program (GP2)

## Abstract

Variants in *PGLYRP2*, particularly rs892145-T, have been suggested as Parkinson’s disease (PD) risk factors. We analyzed data from 48,458 PD patients and 24,670 controls across diverse ancestries. Contrary to previous findings on smaller datasets, no significant sex-dependent effect of rs892145-T was observed across all ancestries. Gene-based analyses identified another variant, rs959117-T, as nominally associated with PD in AAC females (OR=0.65, 95%CI:0.50-0.85, p=1.70E-03), but it did not survive correction for multiple testing (Bonferroni p=0.10). Overall, our results show no association between *PGLRYP2* and PD.

**Highlights:**

- No significant association was found in burden analysis between PGLYRP2 and PD.
- Sex-stratified analysis showed no association between rs892145-T and PD across all ancestries.
- The rs959117-T genetic variant might provide a protective effect against PD in females of African American and Caribbean ancestry.

## 1. Introduction

The etiopathology of Parkinson’s disease (PD) remains unresolved [1]. Studies have implicated the innate immune response in the gut as a potential risk factor for PD [2], such as the case of genetic variants in peptidoglycan recognition proteins (PGRPs) genes [3]. Specifically, the PGLYRP2 protein is involved in proinflammatory responses as well as immune-mediated diseases such as rheumatoid arthritis, and tuberculosis, among others [4, 5, 6, 7, 8]. Animal studies have also suggested that PGRPs play a role in the gut microbiota and brain development [9].

The link between the gene *PGLYRP2* and PD is controversial. In 2023, Ran and collaborators conducted a meta-analysis of existing data on the rs892145-T variant (p.Met270Lys, chr19:15475861A>T [GRCh38]), and investigated its association with PD in a Swedish cohort of 508 PD patients and 585 controls. The authors did not find an association between PD risk and rs892145-T [10]. However, an interaction between sex and rs892145-T was described in which the T allele was found to be significantly less prevalent in female PD patients and more prevalent in male patients, compared to controls, suggesting that rs892145-T may be a risk factor for PD in males (odds ratio [OR] = 1.29, 95% CI:1.01-1.64, nominal P = 0.04). Here we aim to provide further evidence of the potential association between *PGLYRP2* and PD risk by comprehensively exploring this gene across diverse ancestries and in a sex-stratified manner.

## 2. Methods

We leveraged genotyping imputed data from the 12th release of the Global Parkinson’s Genetics Program (GP2; https://gp2.org) [11], genotyped on the NeuroBooster Array [12](NBA; v.1.0, Illumina, San Diego, CA). The dataset consisted of 48,458 PD patients and 24,670 controls after excluding related samples, split into nine ancestries: European [EUR], African American [AAC], African [AFR], Ashkenazi Jews [AJ], Admixed American/Latin American [AMR], Central Asian [CAS], East Asian [EAS], Middle Eastern [MDE], and South Asian [SAS] (Supplementary table 1). All analyses were run separately in each ancestry group. Quality control and genetic ancestry prediction was done with GenoTools (https://github.com/GP2code/GenoTools) as previously described [13]. Additionally, variants with a minor allele count<2, minor allele frequency (MAF) <1%, and Hardy-Weinberg Equilibrium (HWE) p-value *≤* 1 × 10^−4^ were excluded. We also utilized whole genome sequencing (WGS) data from the Accelerating Medicines Partnership® - Parkinson Disease (AMP®-PD; https://amp-pd.org/) release 3, including 2,251 unrelated PD patients and 2,835 controls of European descent.

**Table 1:** PGLYRP2 variant rs959117 associated with Parkinson’s disease among females in the GP2 African American and Caribbean ancestry group (AAC)

| Strata | Genotype | PD<br>(N) | Control<br>(N) | Carrier freq.<br>(PD) | Carrier freq.<br>(Control) | A1 | Allele freq.<br>(PD) | Allele freq.<br>(Control) | OR | 95% CI | p-val | Bonf. |
| --- | --- | --- | --- | --- | --- | --- | --- | --- | --- | --- | --- | --- |
| Male | C/C | 87 | 68 | 26.0% | 23.9% | C | 50.0% | 51.1% | 1.00 | Ref. | NA | NA |
|  | C/T | 161 | 155 | 48.1% | 54.4% | T | 50.0% | 48.9% | 1.04 | 0.83-1.32 | 0.72 | 1.0 |
|  | T/T | 87 | 62 | 26.0% | 21.8% |  |  |  |  |  |  |  |
| Female | C/C | 54 | 106 | 26.1% | 20.9% | C | 54.1% | 47.7% | 1.00 | Ref. | NA | NA |
|  | C/T | 116 | 272 | 56.0% | 53.6% | T | 45.9% | 52.3% | 0.65 | 0.50-0.85 | 1.70E-03 | 0.102 |
|  | T/T | 37 | 129 | 17.9% | 25.4% |  |  |  |  |  |  |  |

Abbreviations: A1, Effect allele; Bonf., Bonferroni corrected p-value; CI, Confidence interval; NA, not available; OR, Odds ratio; PD, Parkinson’s disease; Ref., reference.

*PGLYRP2* gene boundaries (chr19:15,468,645-15,479,501 [GRCh38, NCBI]) were extracted using PLINK2.0 [14]. Annotation was performed using ANNOVAR [15] utilizing RefSeq and ClinVar databases (ver. 20170905). Burden tests were assessed for coding, loss of function, potentially functional variants as well as low frequency variants (MAF < 3%) on PD risk using RVTESTS [16]. Sex, age, and the first five genetic principal components (PC1-5) were used as covariates in all analyses.

Association analyses for *PGLYRP2* variants and sex-stratified analyses focused on rs89214510 [10], were performed using logistic regressions with an additive model in PLINK 2.0[14]. Bonferroni correction (Bonf.) was applied, accounting for all variants identified in the gene within each ancestry group independently. Furthermore, we included an interaction test between rs892145-T and sex utilizing R version 4.4.2, adjusting for age and PC1-5. Additionally, allele and genotype frequencies, and HWE (control group) were calculated using PLINK1.9 [17] and PLINK 2.0 [14], respectively.

For each ancestry group, power calculations were done using the Genetic Association Study (GAS) Power Calculator. An additive model was used at a significance level p=0.05 with a disease prevalence of 0.5% at the OR=1.16 (meta-analysis) [10] and the highest reported OR=1.46 [18] (Supplementary table 2 and 3).

## 3. Results

The total number of identified variants in the genotyping imputed data from GP2 within the *PGLYRP2* genomic region ranged from 70 (AJ, 50 intronic variants) to 337 (EUR, 253 intronic variants). In the AMP®-PD WGS data, a total of 150 variants were identified, including 115 intronic, 27 exonic (19 nonsynonymous), and 8 3’-UTR variants. Additional information on the identified variants can be found in Supplementary table 3.

We evaluated the cumulative effect of *PGLYRP2* variants on PD risk using burden analyses. Nominally significant associations were observed in the MDE ancestry group between PD risk and rare variants (N=82; SKAT p=0.013, SKAT-O p=0.03) (Supplementary table 4). However, none of the associations remained significant after multiple testing corrections for the 60 burden tests performed (p < 8.3E-04).

The rs892145 variant, previously reported to be linked to PD, was in HWE in all ancestry groups (Supplementary table 5). In the logistic regression analyses, no statistically significant association was observed for rs892145-T in any of the ancestry groups (Figure 1, Supplementary table 6).

**Figure 1:**
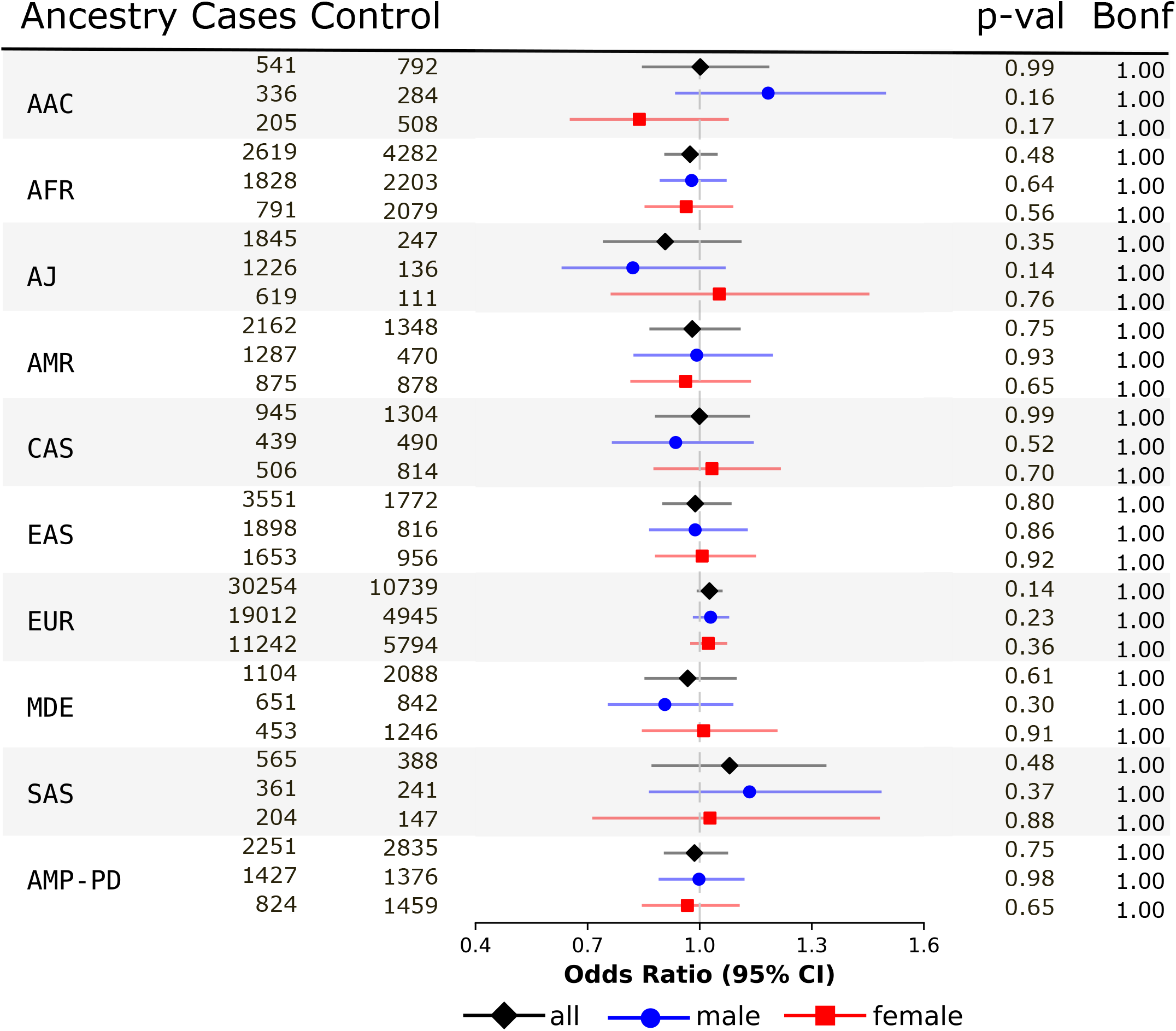
Forest plot showing the association between the *PGLYRP2* rs892145 variant and Parkinson’s disease. Logistic regression analyses using an additive model adjusted by sex, age and PC1-5 for the whole set (black line) or stratified by sex (blue and red lines) in the ancestry groups European [EUR], African American [AAC], African [AFR], Ashkenazi Jews [AJ], Admixed American/Latin American [AMR], Central Asian [CAS], East Asian [EAS], Middle Eastern [MDE], and South Asian [SAS] from the Global Parkinson’s Genetics Program (GP2, release 12) and the dataset from the Accelerating Medicines Partnership - Parkinson Disease (AMP®-PD, release 3).

The interaction test between sex and rs892145-T was only nominally significant in the AAC ancestry group (interaction OR = 1.448, 95% CI: 1.030–2.042, p = 0.034; Supplementary table 7), but the interaction p-value did not survive correction for the ten interaction tests performed (p < 5.0 E-03). The stratified analysis also reflected opposite directions of effect in the two sexes: the T allele trended towards risk in males (OR = 1.183, 95% CI: 0.934–1.499, p = 0.163; Supplementary table 8) and towards protection in females (OR = 0.838, 95% CI: 0.652–1.078, p = 0.169; Supplementary table 9), with neither stratum reaching significance. None of the other ancestry groups showed tendency of a sex-dependent association.

Analysis of all variants in the gene revealed only nominally significant associations (p < 0.05) with PD. The strongest signal arose from rs959117 in the female stratum of the AAC ancestry group (T allele, OR = 0.66, 95% CI: 0.50–0.85, p = 1.70 E-03; Table 1), accompanied by rs7252114 and rs6512037 within a 1.4 kb interval with near-identical genotype counts, indicating a single haplotype. However, none of the associations survived correction for multiple testing.

## 4. Discussion

We investigated the potential relationship between variants in *PGLYRP2* and PD in the largest and most ancestrally diverse dataset to date. No significant results were observed in the whole-gene regression or burden analyses. In contrast with previous studies that showed conflicting results, we did not observe any significant association between *PGLYRP2* variant rs892145-T and PD. No association was reported in the meta-analysis by Ran et al (OR: 1.16, p=0.23) combining previous studies [10]. However, they suggested a potential sex-dependent role for this variant in PD with rs892145-T being described as a risk factor in males (OR = 1.29, 95% CI: 1.01–1.64, p=0.04) with the same direction of effect observed in Chinese and Australian studies [19, 18]. We observed a potential interaction between rs892145-T and sex in the African American and Caribbean (AAC) ancestry group (interaction OR = 1.448, 95% CI: 1.030-2.042, p=0.034) but it did not pass multiple test corrections.

Heterogeneity between the studies of rs892145 in the previously published meta-analysis was observed, with one study reporting that homozygous carriers of the A allele had lower odds of PD (OR = 0.6, 95%CI: 0.40-1.0) in a pooled analysis of American cohorts [3]. However, the minor allele in their study population was the A allele whereas the T allele is reported as the minor allele in all ancestry populations according to the GnomAD browser [20]. Previous positive reports were all based on the heterozygote contrast rather than the additive model and none of the studies corrected for multiple testing, which could explain the discrepancies between previous reports and ours. Fitted here as a sensitivity analysis, the pooled AT-versus-AA odds ratio was 0.995 (0.948-1.045) in males and 1.014 (0.964-1.067) in females, excluding the effects reported by Luan et al. (z = -15.5) and Ran et al. (z = -6.2) [18, 10].

Taking into account the effect size reported by Luan et al. (OR = 1.46)18, the datasets had sufficient statistical power (>0.8) across all ancestries at the observed MAF of 30-40% across populations. Nevertheless, no significant association was observed in EAS in contrast to that described by Luan et al for the Chinese Han population [18]. Using the more conservative effect size reported by Ran et al. (OR = 1.16) [10], only the AFR, AMR, EAS and EUR ancestry groups reached 80% power. Even though the strongest gene-wide signal, rs959117 in AAC females, is independent of rs892145, it did not survive correction (Bonf. p = 0.10) and needs follow-up in future releases. In summary, we find no association between *PGLYRP2* rs892145 and PD in either sex, under either an additive or a heterozygote model.

## Data Availability

All GP2 data is hosted in collaboration with the Accelerating Medicines Partnership in Parkinson's disease and is available via application on the website (https://amp-pd.org/register-for-amp-pd;https://doi.org/10.5281/zenodo.7904832). Data used in the preparation of this article were obtained from the Global Parkinson's Genetics Program (GP2; https://gp2.org). Specifically, we used Tier 2 data from GP2 release 12 (DOI: 10.5281/zenodo.20932193). Tier 1 data can be accessed by completing a form on the Accelerating Medicines Partnership in Parkinson's Disease (AMP-PD) website (https://amp-pd.org/register-for-amp-pd). Tier 2 data access requires approval and a Data Use Agreement signed by your institution. Genotyping imputation, quality control, ancestry prediction, and processing were performed using GenoTools v1.0, publicly available on GitHub (https://github.com/GP2code/GenoTools). All code generated for this article, and the identifiers for all software programs and packages used, are available on GitHub (https://github.com/GP2code/PGLYRP2\_SexDifferences) and were given a persistent identifier via Zenodo (DOI: 10.5281/zenodo.15799510).

https://github.com/GP2code/PGLYRP2_SexDifferences

## Acknowledgements

This work was carried out with the support and guidance of the ‘GP2 Trainee Network’ which is part of the Global Parkinson’s Genetics Program and funded by the Aligning Science Across Parkinson’s (ASAP) initiative. Data used in the preparation of this article were obtained from Global Parkinson’s Genetics Program (GP2). For a complete list of GP2 members, see https://gp2.org. Data used in the preparation of this article were obtained from the Accelerating Medicines Partnership® Parkinson’s Disease (AMP®-PD) Knowledge Platform. For up-to-date information on the study, visit https://www.amp-pd.org. ACCELERATING MEDICINES PARTNERSHIP and AMP are registered service marks of the US Department of Health and Human Services.

## Author contributions: CRediT

**C.L.A**.: Formal analysis, Writing – original draft, Writing – review & editing. **H.M.CS:** Formal analysis, Writing – original draft, Writing – review & editing. **V.Q.P:** Formal analysis, Writing – original draft, Writing – review & editing. **K.A**.: Formal analysis, Writing – review & editing. **K.AB:** Conceptualization, Formal analysis, Writing – original draft, Writing – review & editing.

## Ethical considerations and consent to participate/publication

This study was approved by ethics committees or institutional review boards of all participating sites and conducted in accordance with their ethical standards. Informed consent for study participation was obtained from all participants. Individual study sites participating in GP2 were recruited and samples and data were collected as previously described[21, 22].

## Data and code availability

All GP2 data is hosted in collaboration with the Accelerating Medicines Partnership in Parkinson’s disease and is available via application on the website (https://amp-pd.org/register-for-amp-pd;https://doi.org/10.5281/zenodo.7904832). Data used in the preparation of this article were obtained from the Global Parkinson’s Genetics Program (GP2; https://gp2.org). Specifically, we used Tier 2 data from GP2 release 12 (DOI: 10.5281/zenodo.20932193). Tier 1 data can be accessed by completing a form on the Accelerating Medicines Partnership in Parkinson’s Disease (AMP®-PD) website (https://amp-pd.org/register-for-amp-pd). Tier 2 data access requires approval and a Data Use Agreement signed by your institution. Genotyping imputation, quality control, ancestry prediction, and processing were performed using GenoTools v1.0, publicly available on GitHub (https://github.com/GP2code/GenoTools). All code generated for this article, and the identifiers for all software programs and packages used, are available on GitHub [https://github.com/GP2code/PGLYRP2_SexDifferences] and were given a persistent identifier via Zenodo [DOI: 10.5281/zenodo.15799510].

## References

[1] S. Bandres-Ciga, M. Diez-Fairen, J. J. Kim, A. B. Singleton, Genetics of Parkinson’s disease: An introspection of its journey towards precision medicine, Neurobiology of Disease 137 (2020) 104782. doi:10.1016/j.nbd.2020.104782.

[2] J. Horsager, K. B. Andersen, K. Knudsen, C. Skjærbæk, T. D. Fedorova, N. Okkels, E. Schaeffer, S. K. Bonkat, J. Geday, M. Otto, M. Sommerauer, E. H. Danielsen, E. Bech, J. Kraft, O. L. Munk, S. D. Hansen, N. Pavese, R. Göder, D. J. Brooks, D. Berg, P. Borghammer, Brain-first versus body-first Parkinson’s disease: a multimodal imaging case-control study, Brain: A Journal of Neurology 143 (10) (2020) 3077–3088. doi:10.1093/brain/awaa238.

[3] S. M. Goldman, F. Kamel, G. W. Ross, S. A. Jewell, C. Marras, J. A. Hoppin, D. M. Umbach, G. S. Bhudhikanok, C. Meng, M. Korell, K. Comyns, R. A. Hauser, J. Jankovic, S. A. Factor, S. Bressman, K. E. Lyons, D. P. Sandler, J. W. Langston, C. M. Tanner, Peptidoglycan recognition protein genes and risk of Parkinson’s disease, Movement Disorders: Official Journal of the Movement Disorder Society 29 (9) (2014) 1171–1180. doi:10.1002/mds.25895.

[4] J. Chen, Y.-S. Han, W.-J. Yi, H. Huang, Z.-B. Li, L.-Y. Shi, L.-L. Wei, Y. Yu, T.-T. Jiang, J.-C. Li, Serum sCD14, PGLYRP2 and FGA as potential biomarkers for multidrug-resistant tuberculosis based on data-independent acquisition and targeted proteomics, Journal of Cellular and Molecular Medicine 24 (21) (2020) 12537–12549. doi:10.1111/jcmm.15796.

[5] R. Dziarski, D. Gupta, Review: Mammalian peptidoglycan recognition proteins (PGRPs) in innate immunity, Innate Immunity 16 (3) (2010) 168–174. doi:10.1177/1753425910366059.

[6] F. Huang, X. Liu, Y. Cheng, X. Sun, Y. Li, J. Zhao, D. Cao, Q. Wu, X. Pan, H. Deng, M. Tian, Z. Li, Antibody to peptidoglycan recognition protein (PGLYRP)-2 as a novel biomarker in rheumatoid arthritis, Clinical and Experimental Rheumatology 39 (5) (2021) 988–994. doi:10.55563/clinexprheumatol/vlvlqu.

[7] H. Li, D. Meng, J. Jia, H. Wei, PGLYRP2 as a novel biomarker for the activity and lipid metabolism of systemic lupus erythematosus, Lipids in Health and Disease 20 (1) (2021) 95. doi:10.1186/s12944-021-01515-8.

[8] F. Zulfiqar, I. Hozo, S. Rangarajan, R. A. Mariuzza, R. Dziarski, D. Gupta, Genetic Association of Peptidoglycan Recognition Protein Variants with Inflammatory Bowel Disease, PloS One 8 (6) (2013) e67393. doi:10.1371/journal.pone.0067393.

[9] T. Arentsen, Y. Qian, S. Gkotzis, T. Femenia, T. Wang, K. Udekwu, H. Forssberg, R. Diaz Heijtz, The bacterial peptidoglycan-sensing molecule Pglyrp2 modulates brain development and behavior, Molecular Psychiatry 22 (2) (2017) 257–266. doi:10.1038/mp.2016.182.

[10] C. Ran, K. Wirdefeldt, O. Sydow, P. Svenningsson, R. Diaz Heijtz, Sex Differences in the Allele Distribution of PGLYRP2 Variant rs892145 in Parkinson’s Disease, Parkinson’s Disease 2023 (2023) 6502727. doi:10.1155/2023/6502727.

[11] H. Leonard, M. Nalls, D. Vitale, M. Koretsky, K. Levine, M. Makarious, Z.-H. Fang, L. Jones, E. Voss, J. Taylor, N. Kuznetsov, Global Parkinson’s Genetics Program Data Release 12 (Jul. 2026). doi:10.5281/ZENODO.7904831. URL https://zenodo.org/doi/10.5281/zenodo.7904831

[12] S. Bandres-Ciga, F. Faghri, E. Majounie, M. J. Koretsky, J. Kim, K. S. Levine, H. Leonard, M. B. Makarious, H. Iwaki, P. W. Crea, D. G. Hernandez, S. Arepalli, K. Billingsley, K. Lohmann, C. Klein, S. J. Lubbe, E. Jabbari, P. Saffie-Awad, D. Narendra, A. Reyes-Palomares, J. P. Quinn, C. Schulte, H. R. Morris, B. J. Traynor, S. W. Scholz, H. Houlden, J. Hardy, S. Dumanis, E. Riley, C. Blauwendraat, A. Singleton, M. Nalls, J. Jeff, D. Vitale, Global Parkinson’s Genetics Program (GP2) and the Center for Alzheimer’s and Related Dementias (CARD), NeuroBooster Array: A Genome-Wide Genotyping Platform to Study Neurological Disorders Across Diverse Populations, Movement Disorders: Official Journal of the Movement Disorder Society 39 (11) (2024) 2039–2048. doi:10.1002/mds.29902.

[13] D. Vitale, M. J. Koretsky, N. Kuznetsov, S. Hong, J. Martin, M. James, M. B. Makarious, H. Leonard, H. Iwaki, F. Faghri, C. Blauwendraat, A. B. Singleton, Y. Song, K. Levine, A. A. Kumar-Sreelatha, Z.-H. Fang, M. Nalls, GenoTools: an open-source Python package for efficient genotype data quality control and analysis, G3 15 (1) (2025) jkae268. doi:10.1093/g3journal/jkae268.

[14] C. C. Chang, C. C. Chow, L. C. Tellier, S. Vattikuti, S. M. Purcell, J. J. Lee, Second-generation PLINK: rising to the challenge of larger and richer datasets, GigaScience 4 (2015) 7. doi:10.1186/s13742-015-0047-8.

[15] K. Wang, M. Li, H. Hakonarson, ANNOVAR: functional annotation of genetic variants from high-throughput sequencing data, Nucleic Acids Research 38 (16) (2010) e164. doi:10.1093/nar/gkq603.

[16] X. Zhan, Y. Hu, B. Li, G. R. Abecasis, D. J. Liu, RVTESTS: an efficient and comprehensive tool for rare variant association analysis using sequence data, Bioinformatics 32 (9) (2016) 1423–1426. doi:10.1093/bioinformatics/btw079.

[17] S. Purcell, B. Neale, K. Todd-Brown, L. Thomas, M. A. R. Ferreira, D. Bender, J. Maller, P. Sklar, P. I. W. de Bakker, M. J. Daly, P. C. Sham, PLINK: a tool set for whole-genome association and population-based linkage analyses, American Journal of Human Genetics 81 (3) (2007) 559–575. doi:10.1086/519795.

[18] M. Luan, J. Jin, Y. Wang, X. Li, A. Xie, Association of PGLYRP2 gene polymorphism and sporadic Parkinson’s disease in northern Chinese Han population, Neuroscience Letters 775 (2022) 136547. doi:10.1016/j.neulet.2022.136547.

[19] A. M. Gorecki, M. C. Bakeberg, F. Theunissen, J. E. Kenna, M. E. Hoes, A. L. Pfaff, P. A. Akkari, S. A. Dunlop, S. Kõks, F. L. Mastaglia, R. S. Anderton, Single Nucleotide Polymorphisms Associated With Gut Homeostasis Influence Risk and Age-at-Onset of Parkinson’s Disease, Frontiers in Aging Neuroscience 12 (2020) 603849. doi:10.3389/fnagi.2020.603849.

[20] S. Chen, L. C. Francioli, J. K. Goodrich, R. L. Collins, M. Kanai, Q. Wang, J. Alföldi, N. A. Watts, C. Vittal, L. D. Gauthier, T. Poterba, M. W. Wilson, Y. Tarasova, W. Phu, R. Grant, M. T. Yohannes, Z. Koenig, Y. Farjoun, E. Banks, S. Donnelly, S. Gabriel, N. Gupta, S. Ferriera, C. Tolonen, S. Novod, L. Bergelson, D. Roazen, V. Ruano-Rubio, M. Covarrubias, C. Llanwarne, N. Petrillo, G. Wade, T. Jeandet, R. Munshi, K. Tibbetts, Genome Aggregation Database Consortium, A. O’Donnell-Luria, M. Solomonson, C. Seed, A. R. Martin, M. E. Talkowski, H. L. Rehm, M. J. Daly, G. Tiao, B. M. Neale, D. G. MacArthur, K. J. Karczewski, A genomic mutational constraint map using variation in 76,156 human genomes, Nature 625 (7993) (2024) 92–100. doi:10.1038/s41586-023-06045-0.

[21] L. M. Lange, M. Avenali, M. Ellis, A. Illarionova, I. J. Keller Sarmiento, A.-H. Tan, H. Madoev, C. Galandra, J. Junker, K. Roopnarain, J. Solle, C. Wegel, Z.-H. Fang, P. Heutink, K. R. Kumar, S.-Y. Lim, E. M. Valente, M. Nalls, C. Blauwendraat, A. Singleton, N. Mencacci, K. Lohmann, C. Klein, Global Parkinson’s Genetic Program (GP2), Elucidating causative gene variants in hereditary Parkinson’s disease in the Global Parkinson’s Genetics Program (GP2), NPJ Parkinson’s disease 9 (1) (2023) 100. doi:10.1038/s41531-023-00526-9.

[22] C. Towns, M. Richer, S. Jasaityte, E. J. Stafford, J. Joubert, T. Antar, A. Martinez-Carrasco, M. B. Makarious, B. Casey, D. Vitale, K. Levine, H. Leonard, C. B. Pantazis, L. A. Screven, D. G. Hernandez, C. E. Wegel, J. Solle, M. A. Nalls, C. Blauwendraat, A. B. Singleton, M. M. X. Tan, H. Iwaki, H. R. Morris, Global Parkinson’s Genetics Program (GP2), Defining the causes of sporadic Parkinson’s disease in the global Parkinson’s genetics program (GP2), NPJ Parkinson’s disease 9 (1) (2023) 131. doi:10.1038/s41531-023-00533-w.

